# Estimation of instant case fatality rate of COVID-19 in Wuhan and Hubei based on daily case notification data

**DOI:** 10.1101/2020.03.11.20034215

**Authors:** Lei Cao, Ting-ting Huang, Jun-xia Zhang, Qi Qin, Si-yu Liu, Hui-min Xue, Ya-xin Gong, Chang-hua Ning, Xiao-tong Shen, Jia-xin Yang, Yan-ni Mi, Xue Xiao, Yong-xiao Cao

**Author notes:** Corresponding to. Yong-xiao Cao Ph.D., Professor, Department of Pharmacology, School of Basic Medical Sciences, Xi’an Jiaotong University Health Science Center, Xi’an, Shaanxi 710061, P.R. China, E-mail addresses.

## Abstract

**Background:** The outbreak of coronavirus disease 2019 (COVID-19) initially appeared and has most rapidly spread in Wuhan, China. The case fatality rate is the most direct indicator to assess the hazards of an infectious disease. We aimed to estimate the instant fatality rate and cure rate of COVID-19 in Wuhan City and its affiliated Hubei Province.

**Methods:** We collected the daily case notification data of COVID-19 from Dec 8, 2019 to Mar 10, 2020 in Wuhan City and Hubei Province officially announced by the Chinese authority. The numbers of daily confirmed/deaths/cured cases and the numbers of daily cumulative confirmed/deaths/cured cases were obtained. The death time and cure time of COVID-19 patients were calculated based on the dates of diagnosis, death and discharge of individual cases. Then the estimated diagnosis dates of deaths and cured cases were obtained on the basis of the median death or medium cure time, respectively. Finally, the instant fatality rate of COVID-19 was calculated according to the numbers of deaths and cured cases on the same estimated diagnosis dates.

**Results:** From Jan 1, 2020 to Feb 22, 2020 in Wuhan City, the instant case fatality rate of COVID-19 was 3.4%∼19.5% and the instant cured rate was 80.0%∼96.6%. The average fatality rate reached 11.4% while the average cure rate was 88.6%. During the same period in Hubei Province, the instant case fatality rate was 3.8%∼16.6% and the instant cured rate was 83.4%∼96.6%. The average fatality rate and the average cure rate were 9.2% and 91.8%, respectively.

**Conclusions:** The fatality rate and cure rate of COVID-19 in Wuhan City and Hubei Province were underestimated. Wuhan showed higher fatality rate and cure rate than the whole Hubei Province did.

## Introduction

The novel coronavirus pneumonia (COVID-19)^1^ initially appeared in Wuhan, China is caused by severe acute respiratory syndrome coronavirus 2 (SARS-CoV-2). WHO announced that the ongoing COVID-19 outbreak is a Public Health Emergency of International Concern (PHEIC)^2-4^. As of Mar. 10, 2020, the number of confirmed COVID-19 cases in China has reached to 80955, of which 3162 were dead, and 61567 were cured. Wuhan City and its affiliated Hubei Province are most affected and account for at least 90% of the cases. At present the outbreak in China has been basically controlled. However, the cases of COVID-19 have rapidly increased in other 109 countries around the world, such as South Korea, Italy, Iran, Japan, Singapore, United States and so on. As of Mar 10, 2020, the total number of patients has risen sharply to 32778 outside China, and 876 patients has died.^5^ The global outbreak is getting worse.

Assessing the hazards of infectious diseases is a vital important concern in epidemiology.^6^ The case fatality rate, standing for the percentage of deaths from a disease in the total number of infected-patients during the whole outbreak, is the direct one to reflect the severity of the disease. The accurate case fatality rate of an infectious disease could be obtained after the outbreak is over, on the basis of the numbers of confirmed cases and deaths. Given the severity of COVID-19 outbreak situation throughout the world, it is important to estimate and predict the fatality rate of COVID-19 in the early stage for the hazard evaluation, which might provide the basis for the setting of public security strategy, the allocation of health resources and the adjustment of medical treatment work. It might be also of great help to the government’s decision-making and the public’s understanding.

However, the mortality of COVID-19 published recently in some articles and the “fatality rate” announced by authority is neither mortality nor fatality rate.^7, 8^ Since the numbers of confirmed cases and deaths are constantly changing, it is impossible to get an accurate case fatality rate until the COVID-19 outbreak is over.

We considered that the case fatality rate is a collection from successive instant case fatality rates throughout the disease outbreak. In the present study, we established a method to estimate the instant fatality rate of COVID-19 even the outbreak is still on. We collected the daily case notification data of COVID-19 announced by the Chinese official. The death time and cure time of COVID-19 patients were obtained from individual cases. The estimated diagnosis dates of deaths and cured cases were then calculated. Finally, the instant fatality rate of COVID-19 was estimated according to the numbers of deaths and cured cases on the same estimated diagnosis dates.

## Methods

### Research design

The death time and cure time of COVID-19 patients were calculated based on the dates of diagnosis, death and discharge of individual cases. Then, the estimated diagnosis dates of deaths and the estimated diagnosis dates of cure cases of COVID-19 were respectively calculated on the basis of the median death time or median cure time. Finally, the instant case fatality rate and cure rate of COVID-19 were obtained based on the numbers of deaths and cured cases on the same estimated diagnosis dates.

### Data sources

The daily case notification data of COVID-19 were collected from websites of the National Health Commissions of the People’s Republic of China (http://www.nhc.gov.cn/xcs/xxgzbd/gzbd_index.shtml), Health Commission of Hubei Province (http://wjw.hubei.gov.cn/fbjd/dtyw), and Wuhan Municipal Health Commission (http://wjw.wh.gov.cn/front/web/list3rd/yes/802).

### Data collection

We collected the daily case notification data of COVID-19 in Wuhan City and its affiliated Hubei Province from Dec 8, 2019 to Feb 29, 2020.^9^ The data included the numbers of daily confirmed/deaths/cured cases and the numbers of daily cumulative confirmed/deaths/cured cases. Two researchers collected the data independently every single day, and then checked and corrected the data each other.

### Data analysis

We collected 134 deaths and 536 cured cases. Basing on the dates of diagnosis, death and discharge of individual cases, the death time of COVID-19 patients was the period from diagnosis dates to death dates of the deaths, while the cure time of COVID-19 patients was the period from diagnosis dates to discharge dates of cured cases (figure 1).

**Figure 1.**
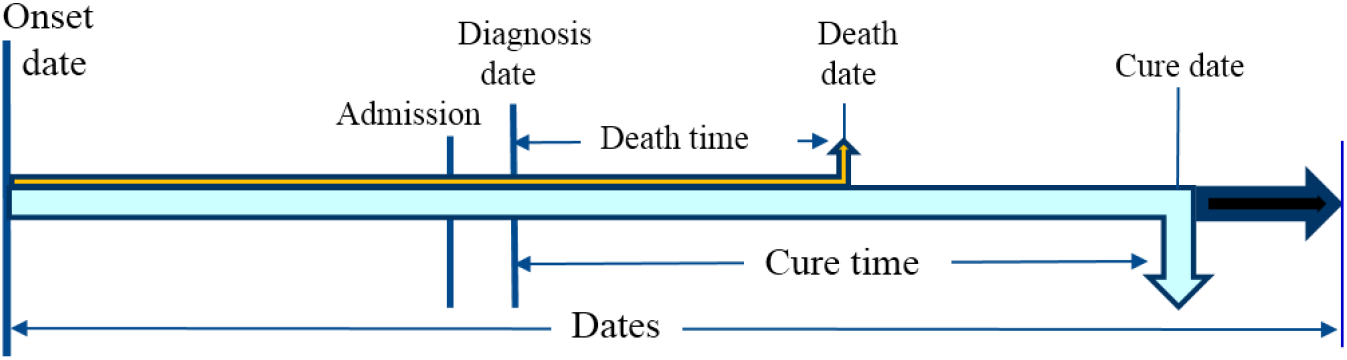
The relationship between death or cure times and dates of diagnosis, death and discharge.

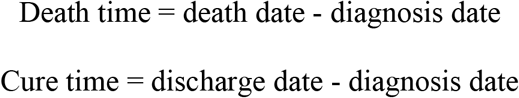

Depending on the above death time and cure time, the diagnosis dates of deaths or cured cases of COVID-19 on daily case notification data were estimated. The estimated diagnosis date (EDD) of deaths is obtained from the date reported on daily case notification data (DCND) of deaths minus mean death time (DT). The estimated diagnostic date (EDD) of cure cases is obtained from the date reported on daily case notification data of cure cases minus mean cure time (CT) (figure 2).

**Figure 2.**
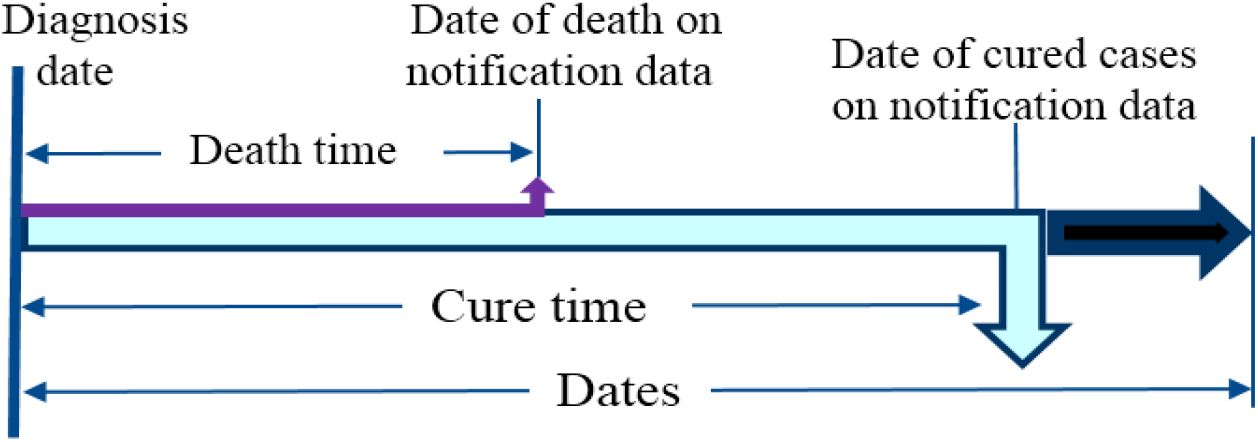
The relationship between estimated diagnosis date (EDD) and dates of deaths or cured cases on daily case notification data (DCND)

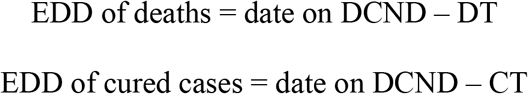

The daily fatality rate (FR) is calculated from the number of cumulative deaths (NCD) divided by a sum of the number of cumulative deaths and the number of cumulative cured cases (NCCC) on the same estimated diagnosis date.

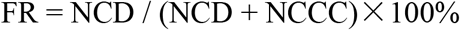

The daily cure rate (CR) is calculated from the number of NCCC divided by a sum of the number of NCD and NCCC on the same estimated diagnosis date.

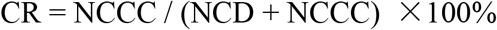

### Statistical analysis

Excel 2016 and GraphPad Prism 8 were used to record, calculate and analyze data.

## Results

### Daily new cases

The curves of daily new COVID-19 cases reported in Wuhan City and Hubei Province were shown in Figure 3. The confirmed cases initially occurred on Dec 8, 2019. The number of daily new confirmed cases started to increase in early January. The number markedly increased in late January, 2020, peaked in mid-February, and then decreased gradually. The death toll began to increase from mid-January to late January, peaked in early February, 2020 and then decreased gradually. The COVID-19 patients were initially cured on Jan10, 2020. The number of daily new cured patients began to increase in mid-January, obviously increased in early and mid-February, still maintained a high level in late February, 2020, and then decreased in the early March. The numbers of daily new confirmed cases, daily new deaths and daily new cured cases in Wuhan City were higher than those in Hubei Province. The characteristics in Hubei Province and Wuhan City were similar.

**Figure 3.**
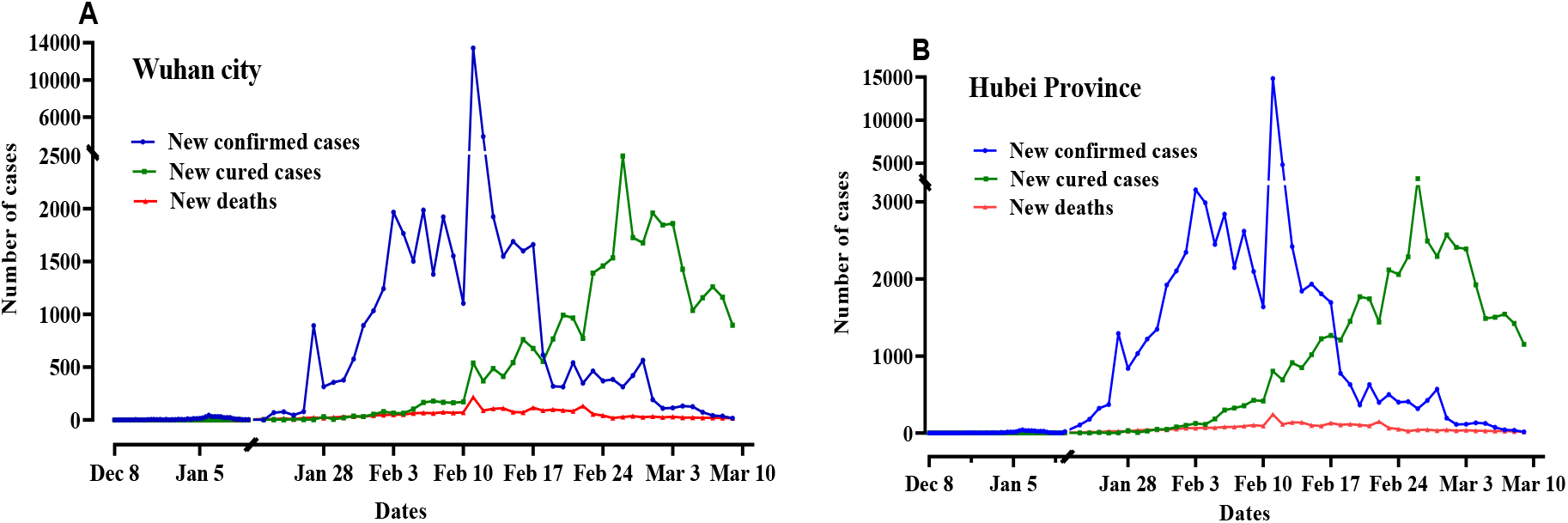
The daily variation of new confirmed cases, new cured cases and new deaths of COVID-19 in Wuhan City (A) and Hubei Province (B).

### Daily cumulative cases

The daily variation curves of cumulative cases of COVID-19 including cumulative confirmed cases, cumulative deaths and cumulative cured cases in Wuhan City and Hubei Province were shown in Figure 4. The number of cumulative confirmed cases began to increase in early January. The increase is sharply accelerated in late January, and stabilized till the early March. The cumulative death toll started to show increase trend in mid-January. The number was remarkably increased in early February, and the number increase slowed down in late February. The number of cumulative cured cases showed an increase tendency in mid-January. The increase was visibly accelerated in early February, and maintain stable till the early March.

**Figure 4.**
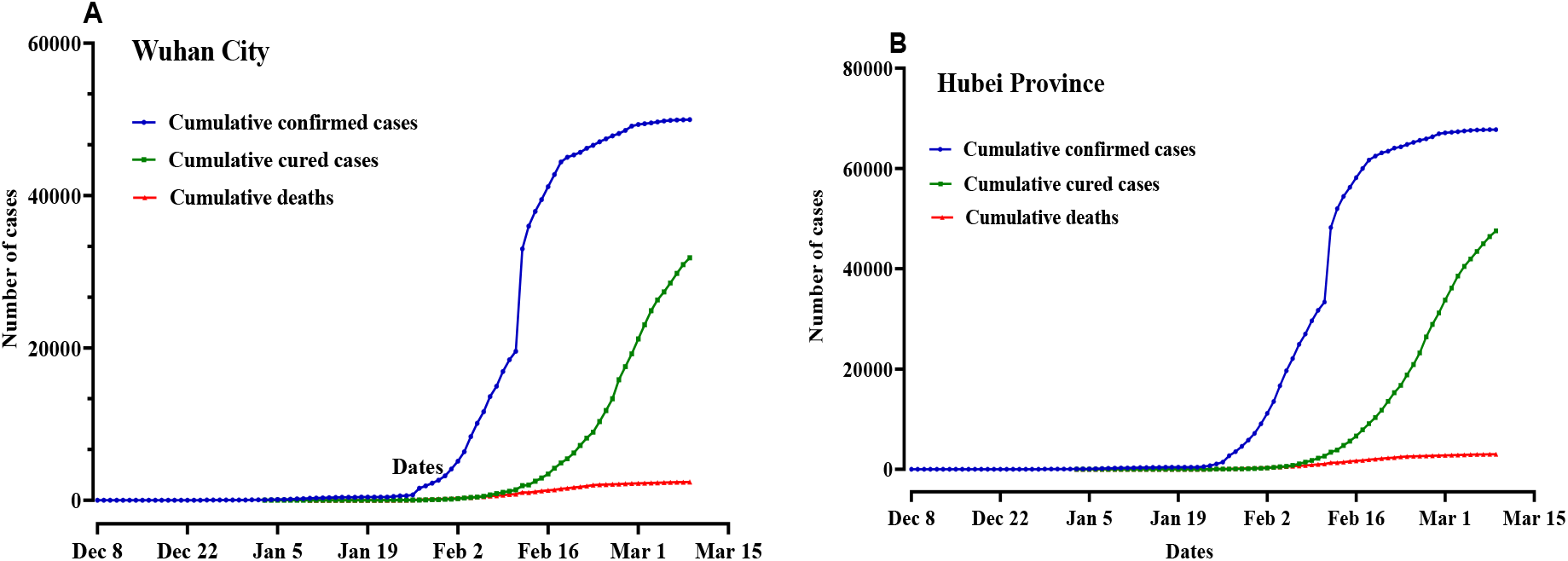
The daily variation of cumulative confirmed cases, cumulative cured cases and cumulative deaths of COVID-19 in Wuhan City and Hubei Province.

### Death time and cure time

After treatment, the COVID-19 patients would end up dead or cured. The time from diagnosis to death is called death time. The time from diagnosis to discharge is cure time. We collected the detailed dates of COVID-19 cases reported on the websites of the National Health Commissions of China and Provincial Health Commissions of China. The death time was calculated on the basis of the diagnosis dates and death dates of 134 dead patients. The mean death time of COVID-19 was 9.2±6.7 days, the median death time was 8 days, and the quartile spacing was 9.2 days. According to the diagnosis dates, and discharge dates of 536 cured cases with COVID-19, the cure time from diagnosis dates to discharge dates was calculated. The cure time curve of the patients displayed a left skewed distribution with a skewness of 0.85. The mean cure time was 13.6±5.6 days, the median cure time of was 12 days, and a 95% confidence interval was from 7.4 to 21.7 (figure 5). Based on the proportion of cumulative confirmed cases who were cured, we estimated that the median cure time of COVID-19 in Wuhan and Hubei was 16 days.

**Figure 5.**
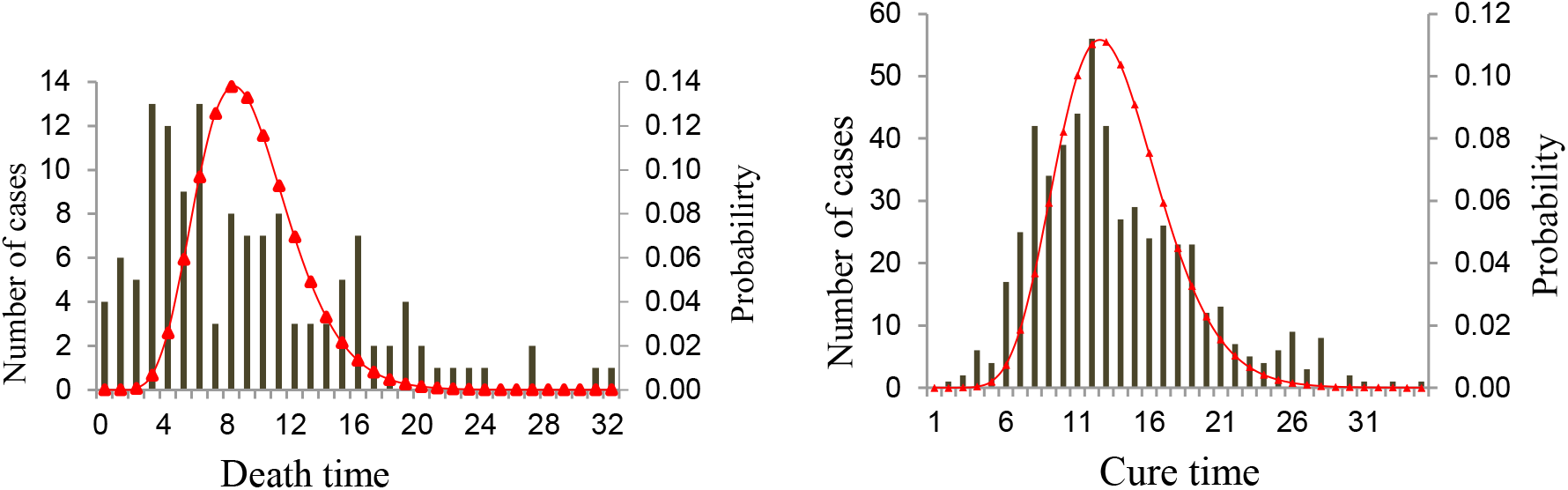
Distributions of death time of 134 deaths and cure time of 536 cured cases of COVID-19.

### Estimation of diagnosis dates of the deaths and cured cases

The confirmed patients would eventually have two endpoints: death or cure. We assumed that the dead patients died on the 8th days, the median death time, after diagnosis. The diagnosis date of the dead patient should be 8 days prior to that date. That is, the estimated diagnosis date (D_d_) of the deaths is the date 8 days before the announced death date (D_i_).

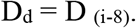

Similarly, we assumed that cured patients are discharged on the 16th days, median cure time, after diagnosis. The estimated diagnosis date of the cure cases is the date 16 days before the announced cure date [D _(i -16)_].

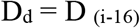

We calculated the diagnosis dates of deaths and cured cases basing on the median death or cure times, respectively. Figure 6 shows the curves of the numbers of cumulative deaths and the number of cumulative cured cases of COVID-19 on the estimated diagnosis dates. The number of confirmed cases was equal to the sum of the number of cumulative cured cases and the number of cumulative death toll on the same estimated diagnosis date. Since the numbers of COVID-19 death toll on the same day was significantly less than the numbers of cured cases, the curve of estimated confirmed COVID-19 cases which was the sum of cumulative cured cases and cumulative death toll, was close to the curve of cumulative cured cases. The shapes and trends of the curves was in Wuhan City are similar to, but lower than those in Hubei province.

**Figure 6.**
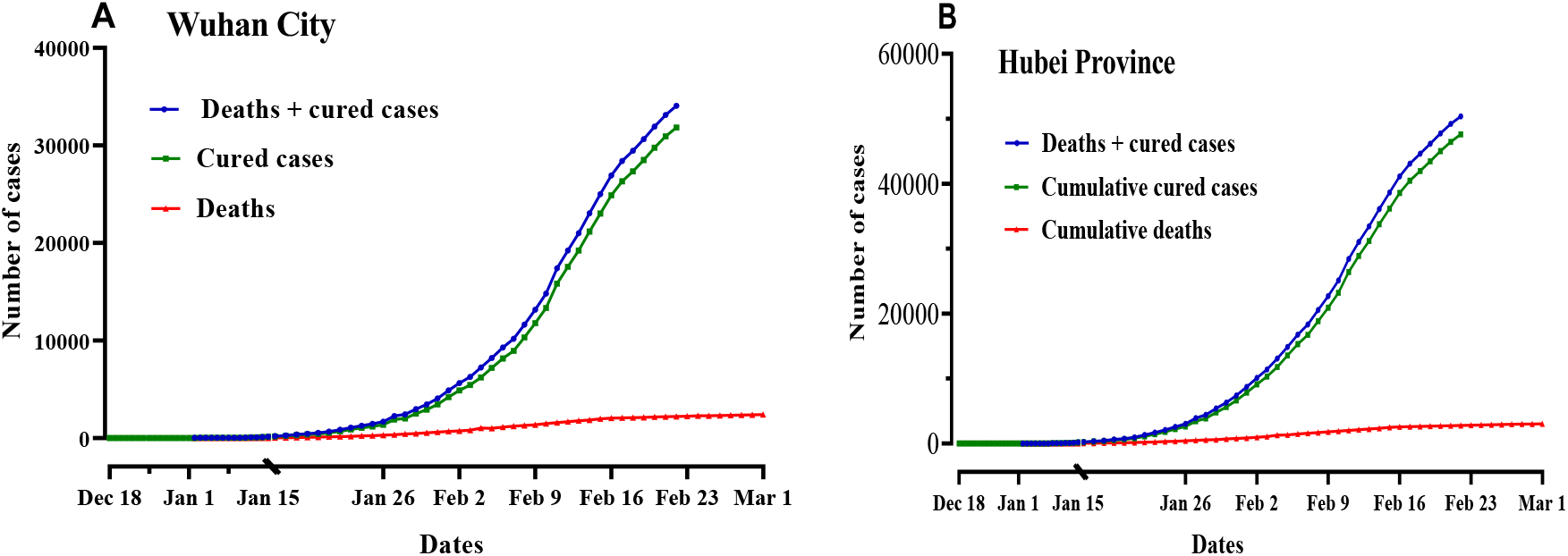
The number of deaths and cured cases on the same estimated diagnosis dates of COVID-19 in Wuhan City (A) and Hubei Province (B).

### Instant fatality rates

The instant fatality rate of COVID-19 in Wuhan City was less than 10% (3.4-9.9%) from Jan 1 to Jan 13, 2020, and gradually increased to 14.2%-19.5% in mid to late January. In February, the instant case fatality rate gradually decreased and stabilized at 6.5% (figure 7A). The average case fatality rate was 11.4%. The instant case fatality rate of COVID-19 in Hubei Province showed the same trend of that in Wuhan City, but the case fatality rates were lower than that in Wuhan City. The instant case fatality rate of COVID-19 in Hubei Province was 10% (3.8-9.1%) from Jan 1 to Jan 13, 2020, then increased to 10.0%-16.6% in late January, and decreased to 5.6% on February 22, 2020 (figure 7B). The average case fatality rate was 9.2%. The curve tendency showed that the case fatality rates were still in decline step by step after February 22, 2020.

**Figure 7.**
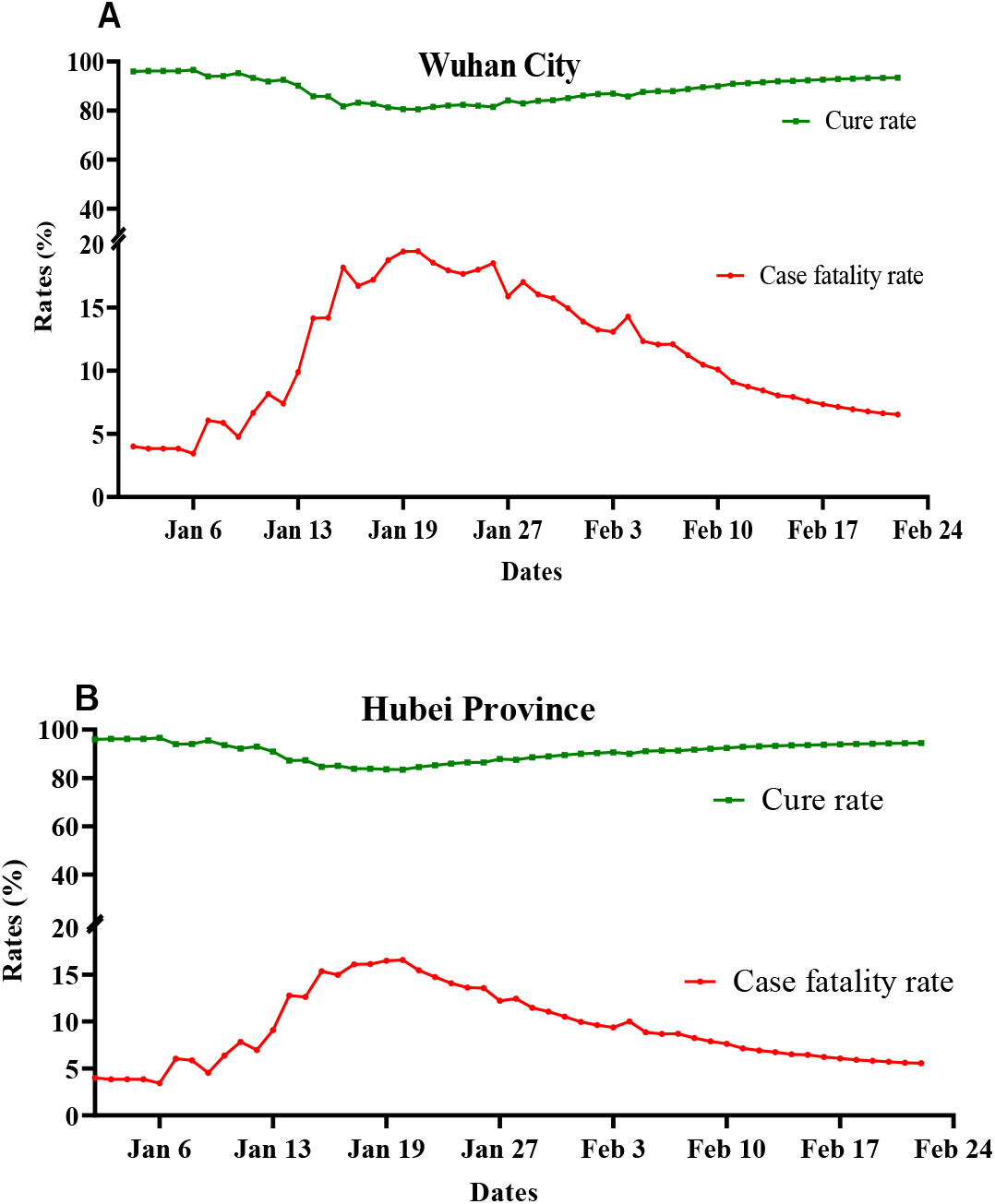
The instant fatality rates and cure rates (B) of COVID-19 in Wuhan City (A) and Hubei Province (B).

### Instant cure rates

The cure rate and case fatality rate are opposite since cure and death are a pair of competing events. The instant cure rate of COVID-19 in Wuhan City was between 80.0%-96.6%, while the rate in Hubei Province was between 83.4%-96.6%. The instant cure rate was lower in second half of January than that in other time. Then the rate gradually increased. On February 22, the instant cure rate increased to 93.5 % in Wuhan, and 94.4% in Hubei (figure 7). The cure rate was likely to continue to increase little by little after February 22, 2020.

## Discussion

The accurate case fatality rate of an infectious disease can be obtained after the outbreak is over. At present, the outbreak of COVID-19 is ongoing throughout China and worldwide. The numbers of confirmed cases, deaths and cured cases are constantly changing.^10^ Therefore, estimating the case fatality rate is obviously challenging. However, it is critical to balance the socioeconomic burden of infection control interventions against their potential benefit for mankind.^8^ The current literatures and the official published case fatality rates have a large deviation. Huang CL *et al* reported clinical features of 41 patients infected with SARS-CoV-2 in Wuhan, China. As of Jan 22, 2020, 28 of 41 patients had been discharged and 6 patients (the mortality 15%) died.^11^ Wang D *et al* reported clinical characteristics of 138 hospitalized patients with 2019 novel coronavirus-infected pneumonia in Wuhan, China.^12^ 47 patients were discharged and 6 died (4.3%). The remaining 85 patients were still hospitalized. The mortality was reported as 4.3%. Zhang YP extracted a total of 72314 cases with COVID-19 reported China’ Infectious Disease information System through Feb.11, 2020.^13^ Thereof a total of 1023 deaths occurred among 44672 confirmed cases for an overall case-fatality rate of 2.3%. However, he ignored a fact that there were 4740 discharged cases and 38909 hospitalized cases in confirmed cases. At present, in official websites the case fatality rates were calculated by dividing the number of known deaths by the number of confirmed cases. These data are neither case fatality rate, nor mortality, and might be off by orders of magnitude.^8^ The confirmed cases included deaths, cured, and hospitalized patients. Some of the hospitalized patients may end up dead in future and their data would not participate in the calculation of the case fatality. In addition, diagnosis of viral infection will precede death or recovery by days to weeks. The number of deaths should be compared to the past confirmed case numbers – accounting for this delay increasing the estimate of the case fatality rate.

We assumed that the case fatality rate of the whole outbreak process could be regarded as a collection of many successive instant case fatality rates. The instant case fatality rate is directly related to various factors at the time, and it is easier to analyze various factors and take possible actions to influence the disease progress by knowing this fatality rate in advance. This instant case fatality rate will gradually approach the case fatality rate as time goes on till the outbreak ends gradually. The extreme instant fatality rate can be used as an approximate value of case fatality rate within certain period of time. This approach provides a way to accurately calculate the fatality rate, which is not affected by hospitalized patients.

We set the instant unit of case fatality rate as a day, and calculated the daily case fatality rate. The daily case fatality rate was equal to the daily cumulative number of deaths divided by the number of deaths plus the number of cured patients. The number of daily confirmed cases is equal to the number of deaths plus the number of cured cases. The confirmed COVID-19 patients had to go through a certain period of time, which is the death time or the cure time, and then result in the final outcomes: death or cure. The date of deaths minus death time is the diagnosis date of deaths. Similarly, the cure date minus the cure time is the diagnosis date of cured patients. The daily case fatality rate and cure rates were then calculated using the cumulative number of deaths and cured cases at the same diagnosis date. This rate varies from day to day with disease-related factors.

The outbreak of COVID-19 firstly appeared in Wuhan, where the situation was most serious with the highest case fatality rate of China. The case fatality rate varied over time. In the first half of January, the fatality rate was less than 10% because the outbreak was not very serious and prevalent. It was easy for patients to see a doctor. But till the beginning of February, the number of patients largely increased and it was difficult for patients to receive medical care treatment. The admission and diagnosis of COVID-19 were delayed, the hospitals were crowded out, and medical condition was deteriorated. As a result, the case fatality rate rose sharply to 20%. In February, a large number of domestic medical resources supported Wuhan, and the medical conditions gradually improved. The treatment concept and methods were updated, the medical level was improved, and the treatment procedure became more and more standardized. On the other hand, the virulence of SARS-CoV-2 may decline as the virus passes through the generations. The proportion of severe cases decreased, and the fatality rate decreased to nearly 6.5%. Basing on this trend, we speculate that the case fatality rate would further decline.

The trend in Hubei Province was similar to that in Wuhan, but the case fatality rate was low. The case fatality rate in the first half of January 2020 was less than 10%. From the second half of January to the beginning of February, it rose to 10% ∼16.6%, and then decreased to 5.6% on February 22. The fatality rate was expected to fall down further.

This study has some limitations. The major influencing factors were mean cure time and death time. The death time was calculated based on the 134 death cases, and the cure time was based on 536 discharged cases. They may deviate from reality and affect the results. We suggest that the government discloses cases of infectious diseases for scientific study to share the resources. Accurate cure time and death time was obtained, it may help to accurately calculate the case fatality rate, and to provide a basis for prevention and control of infectious disease.

## Contributors

YXC, and LC were responsible for conceptual design and led the team. TTH,JXZ did the modeling. QQ, SYL, HMX, YXG, CHN, XTS, JXY were responsible for data collection and management. LC and YXC drafted the manuscript. YNM, and XX revised the manuscript. All authors read the manuscript and contributed to editing.

